# Effect of Graft Fixation Angles in Double-Bundle Anterior Cruciate Ligament Reconstruction, a Systematic Review

**DOI:** 10.1101/2023.04.12.23288505

**Authors:** Rohan Phadke, Bo Song

## Abstract

**Context:** Recent reviews have demonstrated a shift towards anatomical double bundle (DB) techniques for anterior cruciate ligament reconstruction (ACLR). However, to our knowledge, there are no reviews that have directly assessed the relationship between bone tunnel graft angle placement in DB ACLR and clinical outcomes.

**Objective:** To investigate the effect of bone tunnel graft angle on clinical outcomes in DB ACLR.

**Data Sources:** We performed a comprehensive search of the databases PubMed, Scopus, and Embase using the search terms (Fixation OR Angle) AND (ACL OR anterior cruciate ligament) AND (double-bundle OR DB OR double bundle) AND (Reconstruction OR surgery OR arthroscopy) across all time periods.

**Study Selection:** Two independent authors performed a systematic review of the literature. After removing duplicates and applying inclusion/exclusion criteria on a number of clinical outcomes and other factors, 5 eligible studies were recruited under PRISMA guidelines from the 1032 titles identified.

**Study Design:** Systematic Review

**Level of Evidence:** 2a

**Data Extraction:** Systematic review of clinical trials and extraction of relevant surgical data.

**Results:** Our data did not support the original hypothesis and rather showed that increasing the PL bundle angle from 0° to 90° leads to better performance on the pivot-shift tests. Having a nonzero AM bundle graft fixation angle, more specifically at or above 20°, produces better performance on the ATT, Tegner, Lysholm and pivot-shift tests when compared to a 0° angle.

**Conclusions:** A PL bundle angle at or above 45° as well as a nonzero AM bundle angle both yielded better clinical outcome scores. However, with respect to a specific combination, there is no consensus on an optimal graft angle combination in DB ACLR currently. Future randomized controlled trials on this topic are recommended to better understand the effects of greater ranges of graft fixation angles on clinical outcome heterogeneity.

## Introduction

The anterior cruciate ligament (ACL) is a dense band of tissue between the femur and the tibia that serves a vital role in maintaining knee stability during activity. By resisting anterior tibial translation over the femur, the ACL helps prevent hyperextension of the knee. The ACL is one of the most commonly injured structures during sports and is made up of two distinct bundles - the anteromedial (AM) and posterolateral (PL) bundle.^1^ Because the ACL has reduced capacity to inherently heal, surgical repair is often necessary for tears.^1^

ACL reconstruction surgery involves removing the damaged ACL and replacing it with a graft.^1^ Due to the double bundle structure, two distinct methods of anterior cruciate ligament reconstruction (ACLR) have been developed. The double-bundle (DB) technique involves the placement of two grafts in the knee, representing both the AM and PL bundles.^1^ In contrast, the single-bundle (SB) techniques reconstruct only the AM bundle.^1^ While SB grafts are more commonly used, DB grafts are thought to contribute to a more anatomically consistent ACLR at the expense of higher costs and complications.^1^

Recent reviews have demonstrated a shift towards anatomical DB techniques in the last ten years with better levels of clinical recovery, however randomized clinical trials (RCT) comparing SB and DB grafts are still inconclusive.^1,2,3^

Clinical outcomes for ACLR are typically measured by pain and function scores. These include: International Knee Documentation Committee scores (IKDC) for function and symptoms, pivot-shift test for rotary instability, Lysholm scores for symptoms, KT arthrometer scores for instability, range of motion measurements, and radiographic changes.^4^

Fixation devices such as bioabsorbable screws or Endobuttons are used to provide ideal anchoring of the ligament, while portal techniques create the tunnels by which grafts are inserted and vary in position, angle, and width. ^5^ The direct effects of graft fixation angles on clinical outcomes in double-bundle procedures are often understated in clinical trials however they can play a large role in accuracy of graft placement and degree of patient recovery. The different types of graft fixation devices used in surgery range from compression, expansion, or suspension types.^6^ These are further divided by specific mechanisms such as interference screws, cross pin systems, and buttons.^6^ The goal of these devices is to compress or suspend the graft against the surface of the bone or bone tunnel.^6^ The angle with which they are inserted has been shown to have effects in other DB reconstruction procedures. Kennedy et al (2014) found that changing the graft fixation angle combinations of the two grafts being inserted considerably varied knee kinematics compared with both angles being at 0°. ^7^

To our knowledge, there are no reviews that have directly assessed the relationship between bone tunnel graft angle placement in DB ACLR and clinical outcomes. This study aims to address these gaps by performing a systematic review of the literature and comparing clinical outcomes among different trials studying various fixation angles in DB ACLR. Research on the posterior cruciate ligament (PCL) found that increasing posterior bundle fixation angle from 0° corresponded with gradual overloading of the graft.^7^ Furthermore, at anterior bundle fixation angles below 75°, graft forces were more abundant, causing graft attenuation or failure over time.^7^ (Figure 1) Therefore, we hypothesized that a graft fixation combination of the PL graft at 0° and AM bundle graft at either 90° or 105° will result in better clinical outcomes due to reduced strain on the graft.^7^

**Figure 1.**
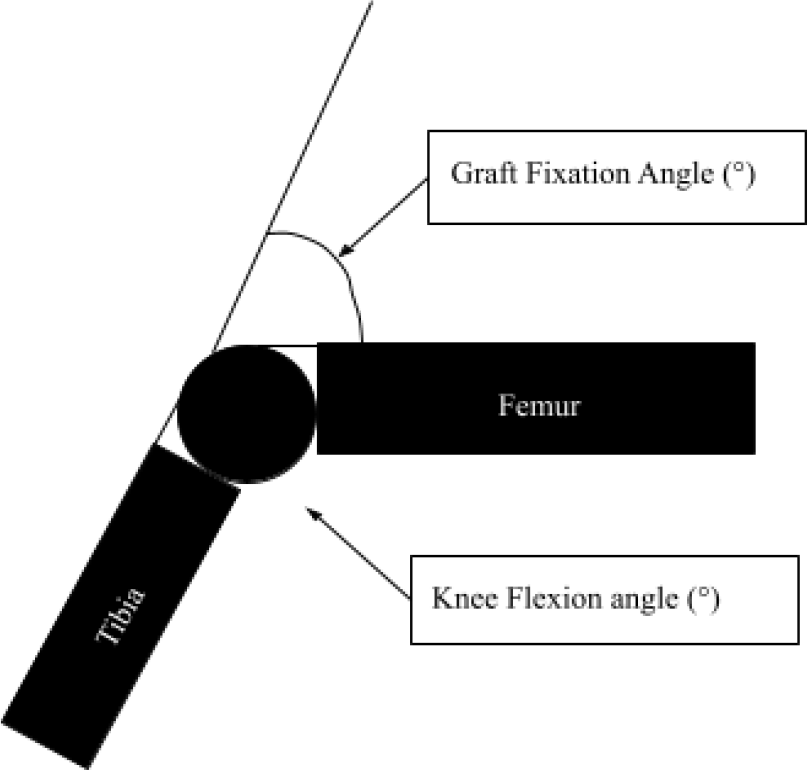
Depicts the graft fixation and knee flexion angles with reference to the femur.

## Methods

Two independent authors performed a comprehensive search of the literature using the databases PubMed, Scopus, and Embase using the search terms (Fixation OR Angle) AND (ACL OR anterior cruciate ligament) AND (double-bundle OR DB OR double bundle) AND (Reconstruction OR surgery OR arthroscopy). After removing duplicates and applying inclusion/exclusion criteria, 5 eligible studies ^8,9,10,11,12^ were recruited under PRISMA guidelines (Figure 2).^13^ A third author was available for any discrepancies in study selection, of which there were none. Given the heterogeneous reporting of data between trials, a comprehensive meta-analysis could not be performed; rather, descriptive statistics were used where possible with measures of variance. Each respective study was analyzed for relevant surgical methodology data and outcome measures.

**Table 1.**
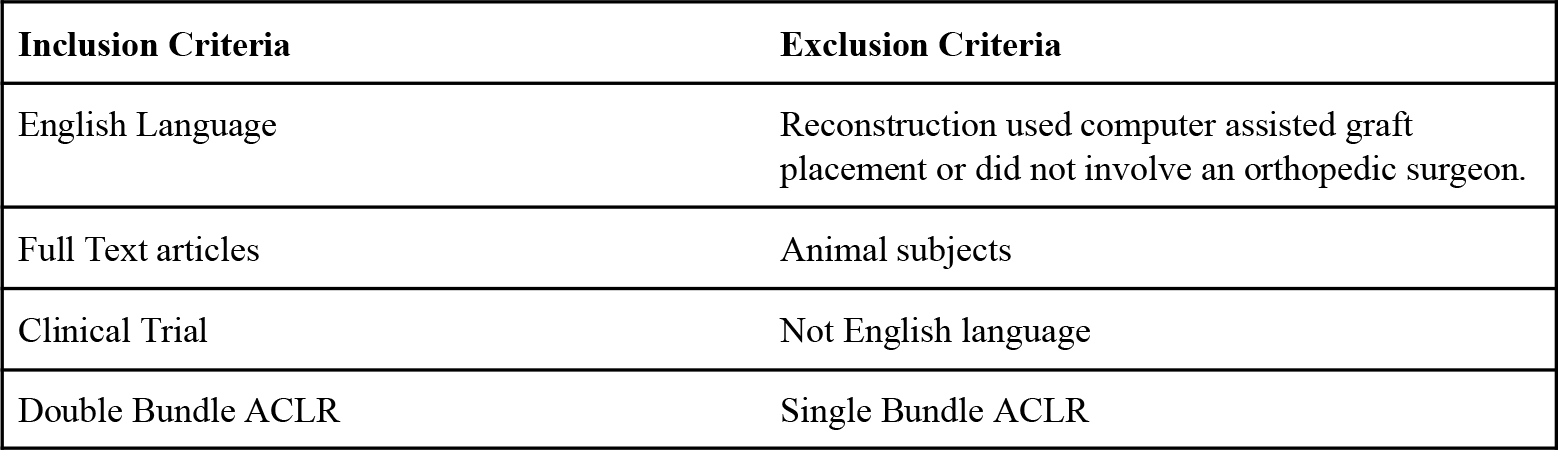
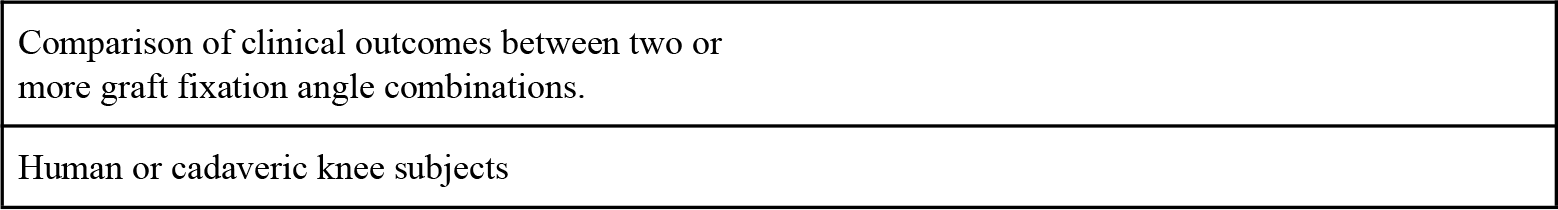
Inclusion and exclusion criteria and corresponding number of studies.

**Figure 2.**
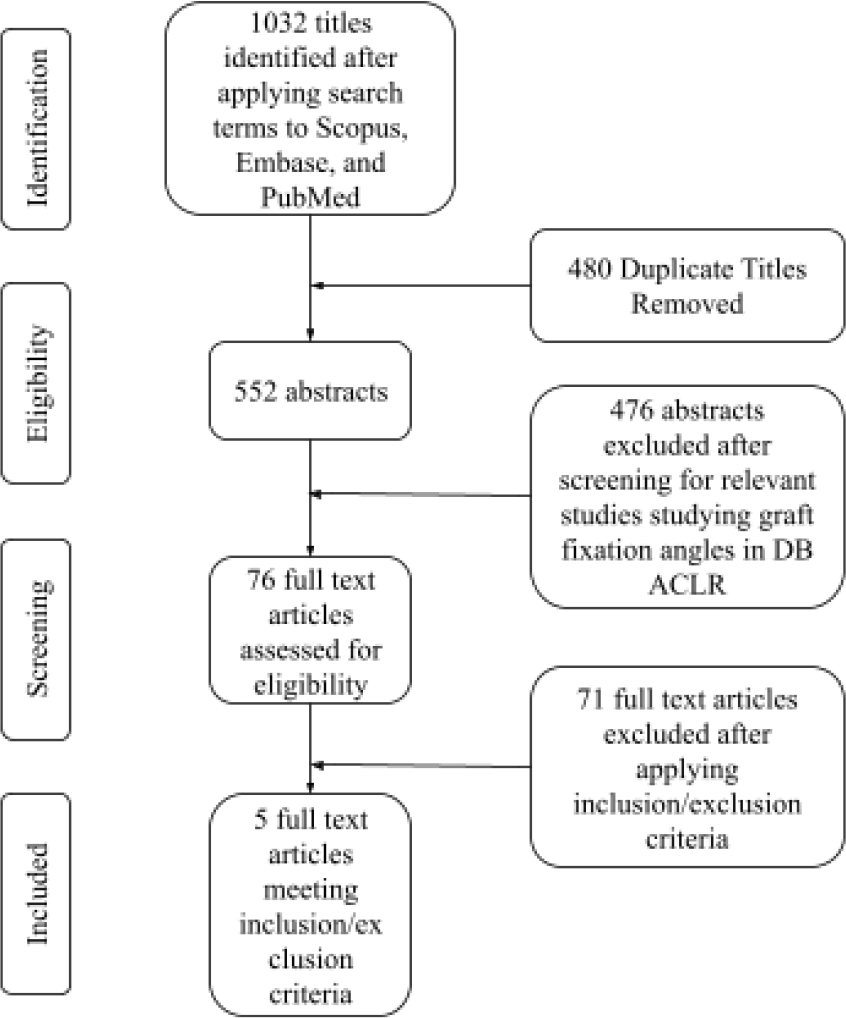
Flow chart depicting the database search for relevant studies.

For inclusion, clinical trials were required to evaluate at least one clinical outcome between two or more graft fixation settings for a sample of human participants or cadaveric knees. Only trials investigating DB ACLR surgeries were included.

Clinical outcome scores were extracted from each trial, specifically data from stability tests, pivot-shift tests, and subjective functional scores. The scores for the tests in each angle cohort were grouped and analyzed for measures of variance such as mean and range.

## Results

### Systematic Review

**Table 2.**
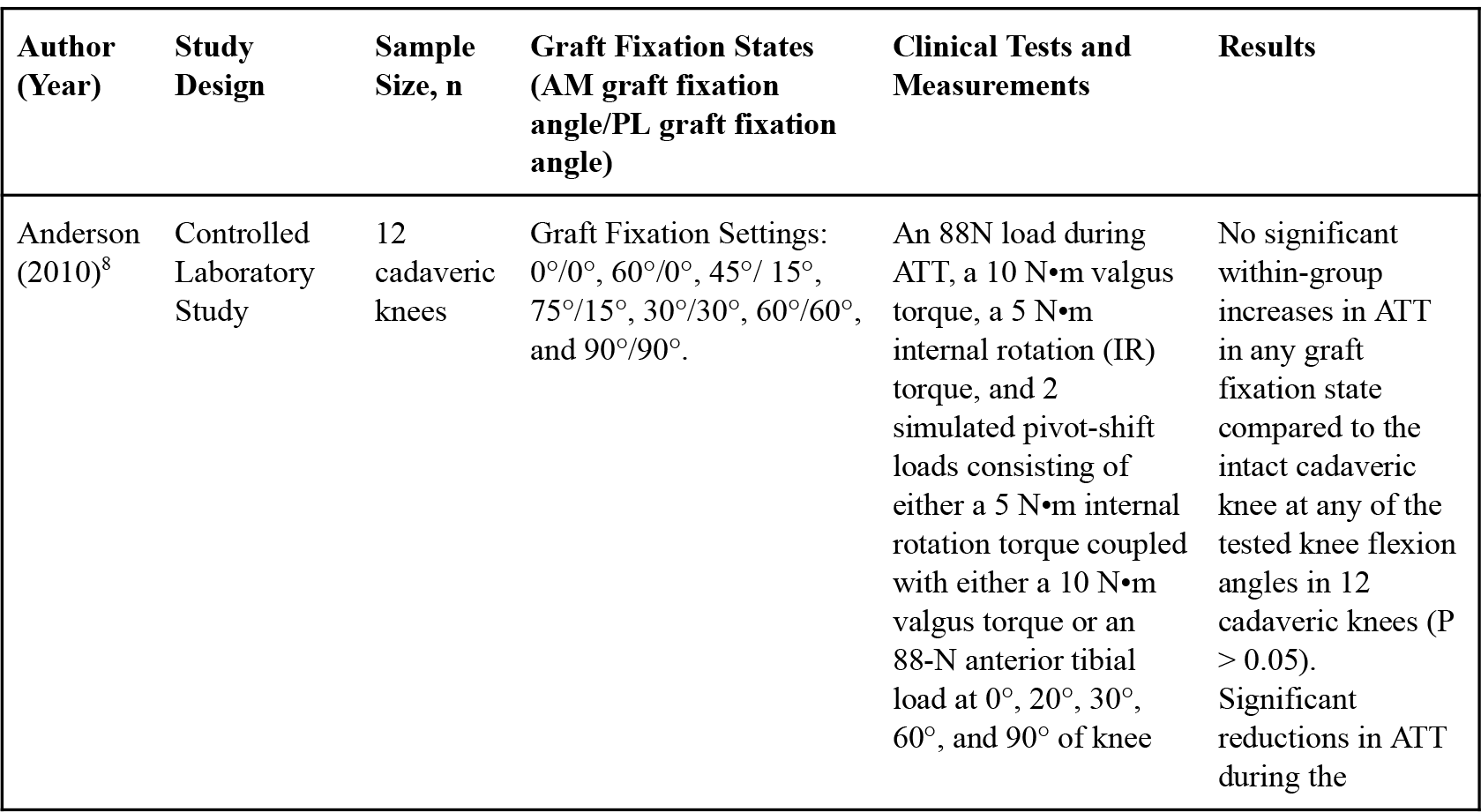
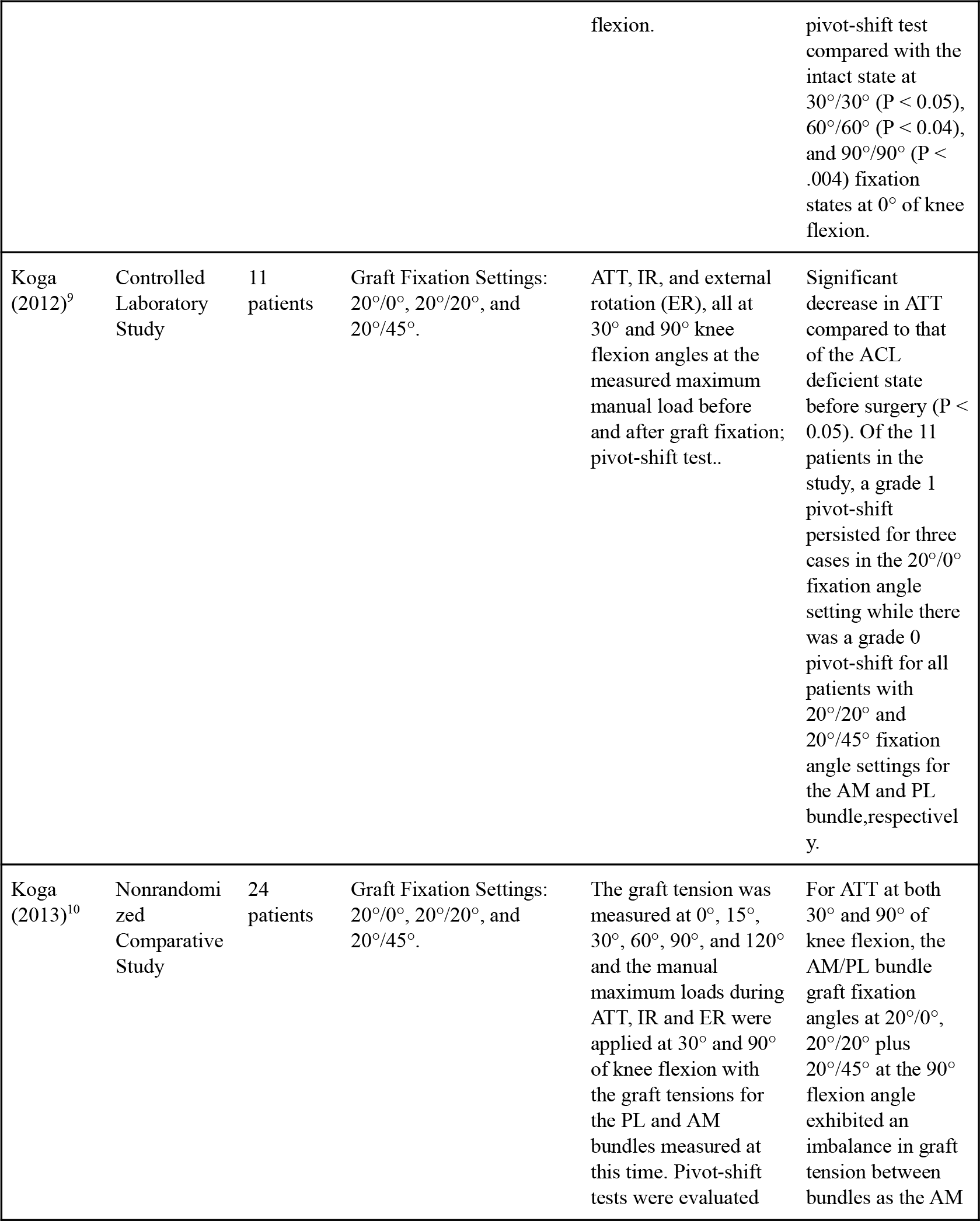
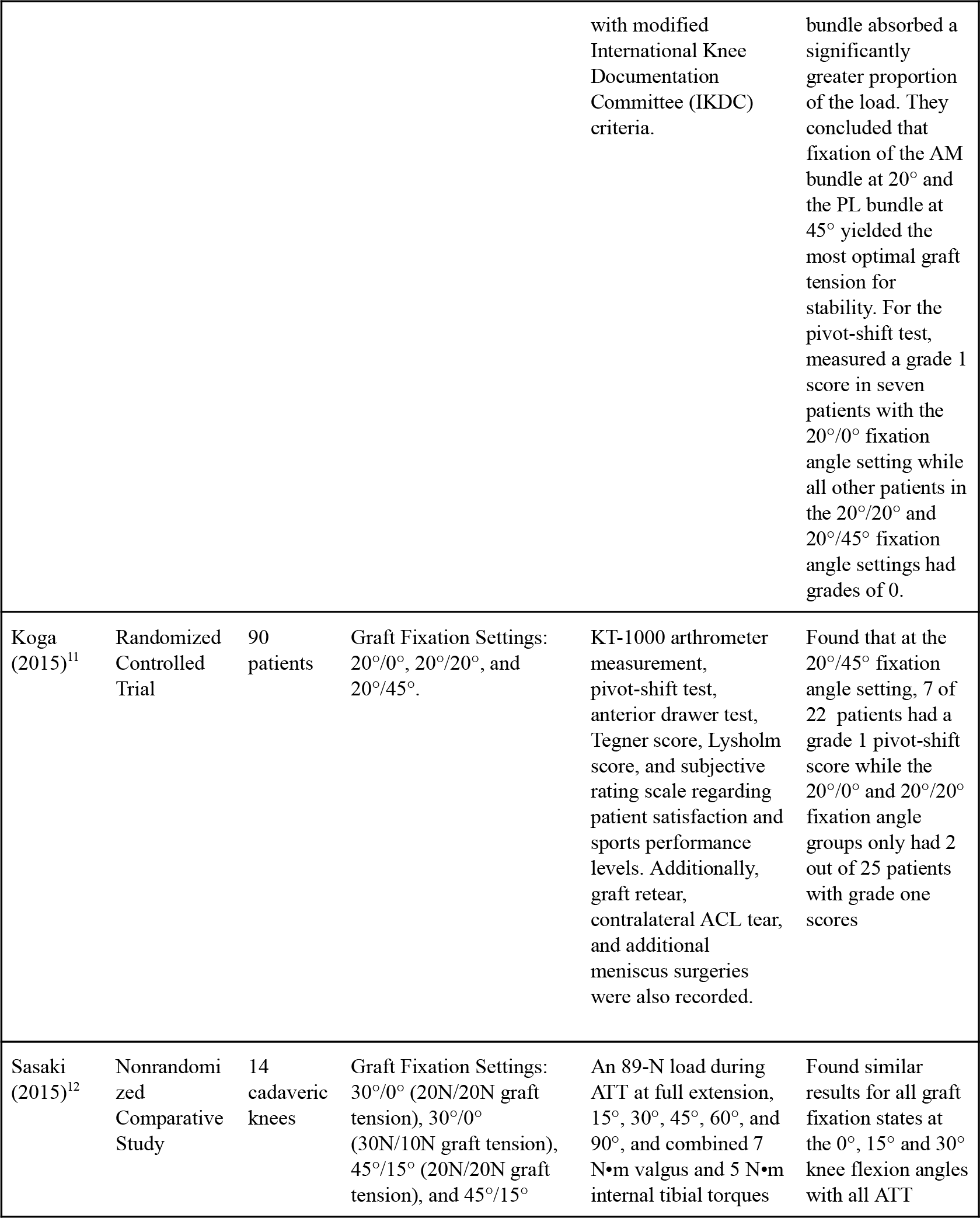
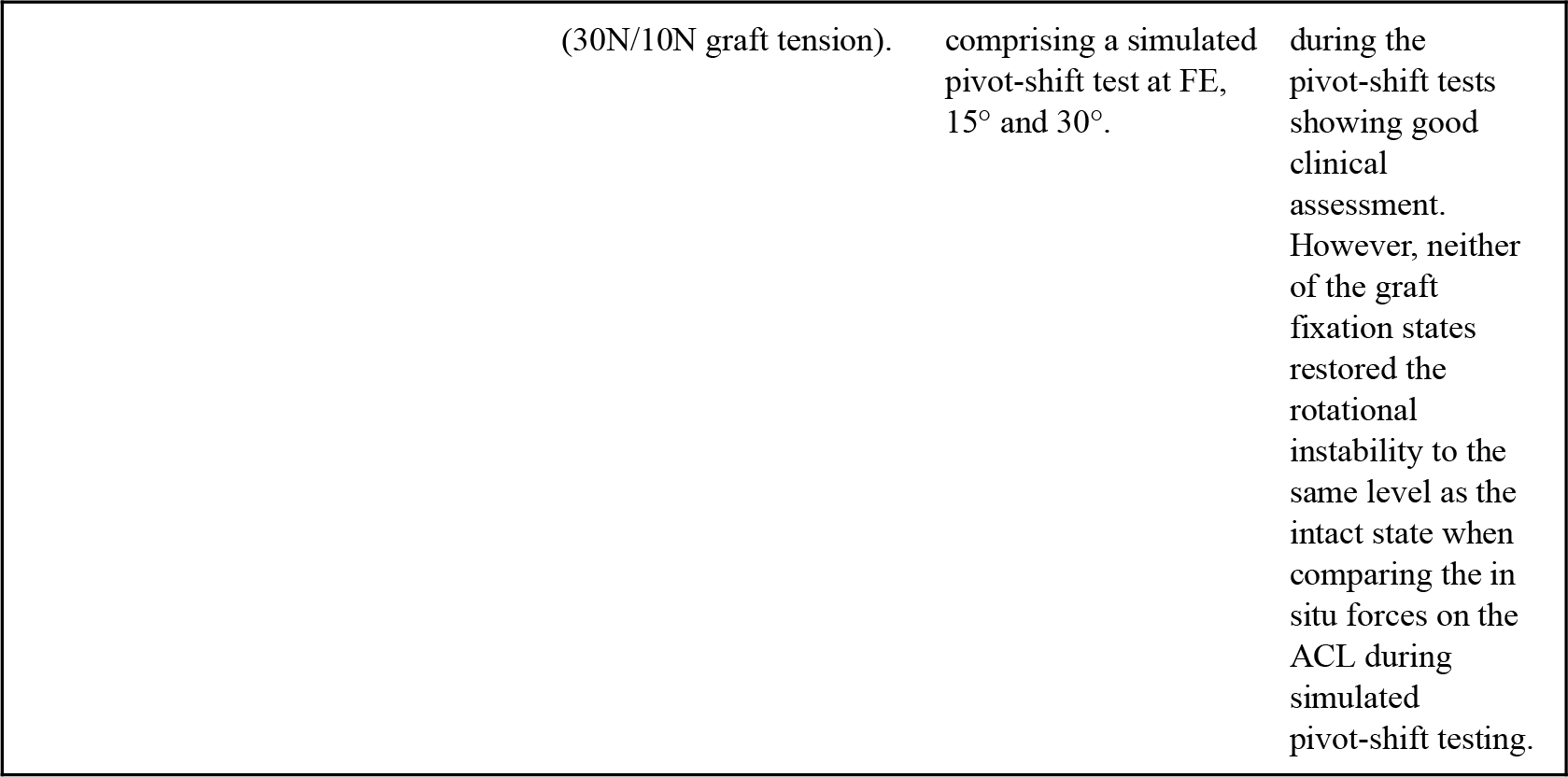
Study characteristics and methodology.

There was a total of 125 patients and 26 cadaveric knees in the 5 included studies. Two of the studies^8,12^ utilized fresh-frozen cadaveric knees. Each trial utilized predetermined graft fixation angle settings for the AM and PL bundles and collected data from certain clinical outcomes. All subjects in this review underwent DB ACLR. Two of the three in vivo studies employed a semitendinosus tendon autograft and an AM portal technique.^9,10^ The third differed in using transtibial tunneling approach for both the AM and PL bundle tunnel drilling.^11^

### Effects of graft fixation angles on anterior tibial translation

ATT represents the anterior movement of the ACL and correlates with the degree of instability. Three studies reported ATT measurements at predetermined graft fixation angles.^8,9,12^ The cadaveric knee studies^8,12^ used the preoperative intact knee as a basis of comparison while the in vivo patient study^9^ used the ACL deficient state. There was no significant difference with respect to the ATT of all the graft fixation states when compared to the intact knee for the cadaveric studies. However, there was a significant difference between all graft fixation settings compared with the state prior to graft insertion in ACL deficient knees. Anderson et al. found no significant within-group increases in ATT in any graft fixation state compared to the intact cadaveric knee at any of the tested knee flexion angles in 12 cadaveric knees (P > 0.05).^8^ On the other hand, Koga et al. (2012) found a significant decrease in ATT compared to that of the ACL deficient state before surgery (P < 0.05).^9^ Koga et al. (2013) recorded tension on the AM and PL bundle grafts in response to ATT, highlighting the constraint levels of the grafts while they resist anterior loads.^10^ For ATT at both 30° and 90° of knee flexion, the AM/PL bundle graft fixation angles at 20°/0°, 20°/20° plus 20°/45° at the 90° flexion angle exhibited an imbalance in graft tension between bundles as the AM bundle absorbed a significantly greater proportion of the load.^10^ They concluded that fixation of the AM bundle at 20° and the PL bundle at 45° yielded the most optimal graft tension for stability.^10^

### Effects of graft fixation angles on pivot-shift rotation

The pivot-shift test is useful for dynamically evaluating for ligamentous laxity to diagnose rotary instability after an ACL rupture but differs from the traditional ATT test in that it measures the ATT after a rotational pivot.^14^ It is scored on a modified IKDC pivot-shift grade scale from 1 to 6 with higher grades correlating to increased rotational instability.^14^ All 5 studies utilized a pivot-shift test as an outcome measure. Anderson et al. found significant reductions in ATT during the pivot-shift test compared with the intact state at 30°/30° (P < 0.05), 60°/60° (P < 0.04), and 90°/90° (P < .004) fixation states at 0° of knee flexion.^8^ At 20° and 30° of knee flexion, there were significant reductions in ATT during the pivot-shift test compared with the intact state at 60°/60° (P < 0.02) and 90°/90° (P < .0003) fixation states.^8^ Of the 11 patients in the study by Koga et al. (2012), a grade 1 pivot-shift persisted for three cases in the 20°/0° fixation angle setting while there was a grade 0 pivot-shift for all patients with 20°/20° and 20°/45° fixation angle settings for the AM and PL bundle, respectively.^9^ Given baseline grades of 3 or 4 in all patients preoperatively, there was a significant overall increase in stability after DB ACLR for any fixation setting.^9^ Koga et al. (2013) measured a grade 1 score in seven patients with the 20°/0° fixation angle setting while all other patients in the 20°/20° and 20°/45° fixation angle settings had grades of 0.^10^ In contrast, Koga et al. (2015) found that at the 20°/45° fixation angle setting, 7 of 22 patients had a grade 1 pivot-shift score while the 20°/0° and 20°/20° fixation angle groups only had 2 out of 25 patients with grade one scores^11^ Sasaki et al. (2015) found similar results for all graft fixation states at the 0°, 15° and 30° knee flexion angles with all ATT during the pivot-shift tests showing good clinical assessment.^12^ However, neither of the graft fixation states restored the rotational instability to the same level as the intact state when comparing the in situ forces on the ACL during simulated pivot-shift testing.^12^

### Effects of graft fixation angles on functional outcomes

The Tegner score is a scale from 1-10 that grades activity based on physical capabilities.^11^ The Lysholm score varies in that it is a 100-point scale that examines specific knee symptoms such as swelling, mechanical locking, or pain while squatting or climbing stairs.^11^ For both tests, higher scores represent better functional ability and fewer symptoms, respectively. Koga et al. (2015) assessed the Tegner and Lysholm scores for 90 patients and found no significant differences in postoperative Tegner and Lysholm scores between the three graft fixation states at 20°/0°, 20°/20°, and 20°/45°.^11^

## Discussion

The objective of this study is to investigate the effects of varying graft fixation angles between the AM and PL bundles in DB ACLR on functional outcomes such as ATT, pivot-shift tests, simulated pivot-shift tests, Tegner scores, and the Lysholm scores.

The literature supports fixation of the AM bundle at and the PL bundle at 20°/45° in yielding a decrease in ATT compared to the ruptured ACL state. This specific graft tensioning is more evenly displaced between bundles which helps prevent over constraint of a single bundle.^10^ Thus, when the AM angle is held at 20°, increasing the PL bundle angle between 0° and 45° yields gradual benefits in graft stability during ATT.^10^ Anderson et al. reported that PL bundle fixation angles of greater than 30° over constrained the knee.^8^ However, Koga et al. (2012) and Koga et al. (2013) found superiority of the 45° PL bundle with regards to ATT.^9,10^ An explanation for this difference could be the use of cadaveric knees in Anderson et al. and the use of a larger range of PL bundle fixation angles up to 90° instead of 45° which allowed for a larger trend to be shown as more data was available.^8^

Analyzing the pivot-shift clinical outcomes at 0°, 20°, and 30° of knee flexion revealed that the 60°/60° and 90°/90° fixation angles were the only combinations that had significant reductions in knee translation compared to the intact state.^8^ Increasing the PL bundle graft fixation angle correlated with improved pivot-shift test scores in cadaveric knees. The increase from 0° to 90° improved performance on the pivot-shift test in the study and thus increased knee stability and the chance at successful reconstruction.^8^ Koga et al. (2012) and Koga et al. (2013) identified gradual pivot-shift grade improvements on a smaller spectrum between 0° and 45° for the PL bundle.^9,10^ An explanation for this trend is that the larger angle caused less ligamentous laxity during the rotational shift.^9,10^ Having a nonzero AM bundle angle was also found to reflect decreased translation during pivot-shift loads in cadaveric knees as evidenced by the clinical results in the 60°/60° and 90°/90° fixation angles by Anderson et al.^8^ The 0°/0° fixation angle combination differed from the hypothesis in Anderson et al. as it did not under constrain the knee due to the lower angle but in fact restored near normal knee stability only without normal tensioning between bundles.^8^ This lack of similarity with native tensioning possibly caused overall inferior stability when compared to the 60°/60° and 90°/90° fixation angle combinations.

When examining subjective scores, the 20°/45° fixation angle combination produced the best results for the Tegner score and the second-best results for the Lysholm score.^11^

Graft tension is defined as the amount of tension each graft exhibits when under pressure during movement.^15^ A study by Murray et al. (2010) that focused primarily on the knee flexion angles and bundle tensions identified the 20°/20° graft fixation setting as the most optimal tensioning setting at five flexion angles tested.^15^ Although this study does not address any specific clinical outcomes, it supports the theory that ATT can be reduced by more optimal graft tensioning. Koga et al. (2013) also addressed graft tensioning but included more graft fixation settings.^10,15^ Their results also differed from those of Murray et al. (2010) in finding that the 20°/45° setting was superior to the 20°/20° setting in graft tensioning during knee flexion.^10,15^ Murray et al. (2010) used only the 20°/20° and a 45°/15° graft fixation settings and no setting with a PL bundle angle of 45°.^15^ Thus, the highest PL bundle angle in the 20°/20° setting for the study may explain the superior graft tensioning.^15^ Another study by Luites et al. only incorporated one graft setting fixed at a 90°/20° knee flexion angle; this study showed no significantly difference in anterior laxity at all flexion angles after DB ACLR.^16^ Sasaki et al. (2015) found with regards to the AM bundle angle that fixation at a lower angle can indicate better rotational function as seen through simulated pivot-shift tests.^12,17,18^ Therefore, a nonzero AM bundle fixation angle that is also below 45° may provide superior clinical outcomes to greater angles as supported by the literature.

The original hypothesis for this study was based on previous research on the PCL and its anatomical similarity to the ACL with reference to graft angles.^7^ Our data does not support the original hypothesis that a 90°/0° or 105°/0° fixation angle combination would produce the best clinical outcomes.^7^ In fact, fixation of the PL bundle at 0° did not demonstrate any statistically significant or superior clinical outcomes compared to other angles. Increasing the PL bundle angle from 0° to 90° leads to better performance on pivot-shift tests.^9, 10^ Koga et al. (2012) and Koga et al. (2013) both reported pivot-shift scores of 0 for the 20° and 45° PL bundles while patients in the 0° PL bundle cohort had scores of 1 for both studies.^9, 10^ One explanation is that the larger PL bundle graft fixation angle may resist rotational movement more efficiently. Similarly, having a nonzero AM bundle graft fixation angle has been proven to reflect good performance on ATT, Tegner, Lysholm and pivot-shift results when compared to a 0° angle.^11^ Based on the limited fixation angle combinations present in this study, a 20°/45° combination showed statistically significant improvements in ATT, pivot-shift tests, Lysholm and Tegner scores when compared to the ACL deficient state.^9, 10, 11^

One notable limitation of this study was the reduced scope due to the low number of trials on this topic. Furthermore, there was significant heterogeneity in the data especially with regards to study design and methods, with some utilizing cadaveric knees and others using human patients. Cadaveric knees may exhibit differences with respect to bone quality and ligament properties.^3^ Specifically, there were some prominent differences between the placement and number of the femoral and tibial tunnels as well as the amount of graft pretension between the DB ACLR studies. Due to this heterogeneity, a true meta-analysis could not be performed. There were also few studies that included PL bundle graft fixation angles between 45° and 90°. This limited the possibility of conclusions capable of addressing the effects of the full range of fixation angles. Other limitations included variable follow up times and subjective nature of clinical outcome measurements.

Future studies can evaluate clinical outcomes in patients undergoing DB ACLR with varied fixation angles that fit the accepted spectrum for AM and PL bundles. Other clinical outcomes can be studied including Kellgren Lawrence scores, graft failure rates, and revision surgery success rates. Ideally, future studies should also employ randomized clinical trials with long-term follow up to be able to further assess the implications of reconstruction.

## Conclusion

Increasing the PL bundle angle from 0° to 90° leads to better performance on pivot-shift tests. On the other hand, having a nonzero AM bundle graft fixation angle has been proven to reflect good performance on ATT, Tegner, Lysholm and pivot-shift results when compared to a 0° angle. Based on the limited fixation angle combinations present in this study, a 20°/45° combination showed statistically significant improvements in ATT and pivot-shift tests when compared to the ACL deficient state. Future studies are recommended to draw definitive conclusions on which graft angles are most optimal.

## Data Availability

All data produced in the study are available online

## Notes

### Competing Interest Statement

The authors have declared no competing interest.

### Funding Statement

This study did not receive any funding

